# NO_2_ pollution decrease in big cities of Latin America during COVID-19 pandemic

**DOI:** 10.1101/2022.08.08.22277819

**Authors:** Matias Poullain, Juan Martin Guerrieri, Manuel Eduardo Miller, María Eugenia Utgés, María Soledad Santini, Mariana Manteca Acosta, Agustín Fernández, Franco Leonel Marsico

## Abstract

NO_2_ is a mainly anthropogenic gas that affects population health and its exposure is associated with several respiratory diseases. Its tropospheric concentration is associated with vehicle emissions. During 2020, COVID-19 lockdowns have impeded population’s mobility, hence constructing an almost ideal situation to study their relationship with tropospheric NO_2_ concentration. We used TROPOMI (TROPOspheric Monitoring Instrument) satellite images, Google mobility reports and vehicule count in order to study these relationships in six big Latin American metropolitan areas: México DF, São Paulo, Buenos Aires, Rio de Janeiro, Lima and Bogotá. In all of them, tropospheric NO_2_ concentration decreased during 2020 compared to 2019, particularly during April 2020. Temperature differences alone could not explain the NO_2_ concentration differences between February and April 2020. The daily vehicle count in Buenos Aires was a significantly important variable in order to explain NO_2_ concentration variations (p < 0.001) and it could be replaced by the daily Google’s residential variation without significant information loss (p ≃ 1). This study strengthens previous research findings about NO_2_ concentration reduction during COVID-19 lockdowns and shows the relationship between human mobility and air pollution in the particular context of Latin America big cities.

## 1 Introduction

### 1.1 Mobility reduction during COVID-19 pandemic

The current outbreak of the new coronavirus SARS-CoV-2, first reported in Hubei Province of the People’s Republic of China, has spread to many other countries. On January 30^th^, 2020, the WHO Emergency Committee declared a global health emergency based on increasing case reporting rates. During several months of 2020, most countries implemented different strategies such as social distancing measures against the spread and transmission of COVID-19. For instance: imposing lockdowns, banning large gatherings, closing schools, restaurants, etc. These measures proved to be effective in their purpose [1, 2, 3, 4, 5, 6, 7, 8]. Hence, in order to understand population mobility during COVID-19 lockdown, it is necessary to measure it directly. Many researchers have used mobile data and related it to COVID-19 outbreaks [9, 10, 11], socioeconomic consequences [12, 13, 14], and environmental changes [15, 16].

Latin America is a very affected area by COVID-19 outbreak [17]. These cities are: México DF, México (24.5 millions inhabitants), São Paulo, Brasil (22.6 millions inhabitants), Buenos Aires Metropolitan Region, Argentina (16.6 millions inhabitants), Rio de Janeiro, Brasil (13.3 millions inhabitants), Lima, Perú (11.1 millions inhabitants) and Bogotá, Colombia (9.8 millions inhabitants).

Latin American governments’ responses were very different between countries [18, 19], especially at the very beginning of the pandemic. For instance: Argentina, Perú and Colombia have followed WHO recommendations very closely, declaring sanitary emergencies less than 10 days after each detected its first COVID-19 case and implementing total closures shortly after. On the contrary, México and Brasil declared a sanitary emergency nearly a month after their first detected case and only partial closures were implemented. Though, in all mentioned cities, both weak and strong restrictions started in March 2020 [18, 19].

### 1.2 NO_2_ pollution and health

During COVID-19 lockdown, many studies [20, 21, 22, 23, 24, 25, 26, 27] described the improvement of air quality in big cities around the world associated with decreasement of air pollutant emissions during COVID-19 measures implementations. Particularly, nitrogen dioxide (NO_2_) is a common air pollutant and it has natural (as soil emissions or forest fires, for example) and anthropogenic sources. The predominant ones are fossil fuel combustion, biomass burning, livestock waste, biogenic soil and indoor combustions [28, 29], though many studies have described the vehicle transportation as the mainly emisor of NO_2_ and NO_X_ (NO

+ NO_2_, being NO_2_/NO_X_ ratio increasing [30, 31, 32]. A worldwide study [33] states that the highest ground values of NO_2_ where observed in the principal cities of America, Europe and Asia, where the common denominator is the high number of vehicles, which are clearly the main source of NO_2_ emissions in cities worldwide. Moreover, the reduction of the NO_2_ exposure in Latin America requires considerable investments in public transportation, creative traffic management, improving fuel quality, promoting modes of transportation, and expanding low emission zones [34]. Although NO_2_ and NO_X_ emissions are decreasing in Europe associated to the new vehicles and fuels technologies [32], in Argentina, NO_X_ increasing emissions (21% between 1995-2019) are still associated with economic growth [35], where it is estimated that 84% of them are concentrated in urban areas of more than 2.500 inhab./km^2^ and that 83% of all ozone precursors (including NO_2_) are emitted by transport, 4% by the industry, 6% by residence and 6% by electricity production [36].

It is an important atmospheric trace gas due to its health effects and its environmental effects: long term exposure to NO_2_ is associated with an increase in Chronic Obstructive Pulmonary Disease (COPD) and Acute Lower Respiratory Infection (ALRI) mortality [37]. It absorbs visible solar radiation and contributes to impaired atmospheric visibility and has a potentially direct role in global climate change. WHO states [38] that “nitrogen dioxide is subject to extensive further atmospheric transformations”, and that “through the photochemical reaction sequence initiated by solar-radiation-induced activation of nitrogen dioxide, the newly generated pollutants are an important source of organic, nitrate and sulfate particles currently measured as PM_10_ or PM_2.5_. For these reasons, NO_2_ is a key precursor of a range of secondary pollutants whose effects on human health are well-documented”.

This gas has a seasonal behavior due to natural factors involved in its destruction process such as the number of hours of sunlight, air temperature or ozone concentration [38]. Likewise, a seasonal change in the behavior of electricity and fossil fuel consumption is associated with the concentration of NO_2_ in urban centers during winter (due to the increase in electricity consumption associated with domestic heating) being higher than during summer. On the other hand, in winter, low temperatures and the reduction in sunlight hours inhibit the NO_2_ decomposition reaction in which ozone takes part (ozone also has a seasonal behavior that modulates the decomposition of NO_2_ [39]). During this season, thermal inversions are favored and therefore the stability of the planetary boundary layer, which inhibits the mechanical mixing and prolongs the presence of NO_2_ in the atmosphere. The concentration of NO_2_ varies depending on the meteorological conditions on the Earth’s surface. Several studies show that the NO_2_ concentration presents a higher concentration and greater variability in the presence of low wind speeds and little precipitation. Both, wind speed and precipitation, favor the destruction of NO_2_ [40, 41]. Given that NO_2_ emission is associated with traffic, its concentration variability has a weekly pattern, as it is observed in Europe where the emission during weekends is significantly lower [42].

Most of the Latin American big cities are located in developing countries. As previously observed by Mage et al. [43], in these countries air pollution represents a major risk for healthcare due to the lack of resources and difficulties in the delivery of medical care in acute cases. Particularly, in Latin America air pollution is essentially a metropolitan problem. The hyper centralized urban growth led to cities with high traffic congestion, inadequate urban transport infrastructures and an aging motor vehicle fleet [44]. In general, low income neighborhoods are located surrounding the metropolitan areas, leaving the most vulnerable population exposed to poor quality urban air. This shows the relevance of constant and precise monitoring of air pollution and its impact on human health in these cities.

### 1.3 Previous research

Previous works have explored the NO_2_ and human mobility reduction during the COVID-19 pandemic. Several studies have found a strong positive correlation between human mobility and NO_2_ concentration. In different cities of the USA, an average drop of NO_2_ column concentration of 13% during 2020 and a 20% average mobility reduction during March 2020 were observed [45]. Another study has found a greater NO_2_ concentration decrease in cities of USA (at least 23%) to be strongly correlated with a 20% average decrease in urban mobility for workplaces [46]. Further, in Singapore a 54% NO_2_ concentration reduction which is significantly correlated (Spearman) to human mobility reduction [47]. On the other hand, a worldwide study has described a maximum of 70% NO_2_ Air Quality Index (AQI) reduction observed in the Philippines, seconded by Lima, Perú with a 55% and reaching a 60% mobility reduction across Europe, Asia, and the Americas [48, 49].

Nevertheless, there is a lack of comparative studies of the association between these two features in Latin America. In this work, we analyze the largest cities in Latin America through a comparative study approach, in order to analyze the association between tropospheric NO_2_ concentration and population mobility. This study involves obtaining data from open sources and databases allowing its reproducibility. Our main hypothesis is that during the first months of COVID-19 outbreak there was a tropospheric NO_2_ concentration reduction in Latin American large cities. Furthermore, this phenomenon was related to population mobility decrease.

## 2 Methods

### 2.1 Study design and data

The reason for choosing Latin American cities is the hyper centralized urbanization, health care accessibility, economic influence of the region and socio-economical inequalities. Latin American countries are significantly tied with higher consumption of fossil fuel which emits the highest level of GHG emissions than any other economic sector [50]

The data used were taken from a number of open access sources and have different temporal and spatial resolutions. All the information used corresponds to the six most populated metropolitan areas in Latin America (referred as cities unless it’s specified otherwise), which are Buenos Aires Metropolitan Region (Argentina), São Paulo (Brasil), Rio de Janeiro (Brasil), Lima (Perú), México DF (México), and Bogotá (Colombia).

The tropospheric offline NO_2_ dataset (OFFL NO_2_) was extracted from Sentinel-5P, an Earth observation satellite developed by the European Space Agency (ESA) as part of the Copernicus Programme. Its spectrometer TROPOMI (TROPOspheric Monitoring Instrument) measures the concentration of different atmospheric gases such as NO_2_, O_3_ and CH_4_ among others.

L3 data set was downloaded from the Google Earth Engine (GEE) Javascript API and with a resolution of 0.01° × 0.01° as a result of the oversampling of the L2 OFFL NO_2_ data from TROPOMI by GEE [51, 52, 40]. The conversion to L3 is done by the harpconvert tool using the bin spatial operation. GEE OFFL NO_2_ data set (in units of *µ*mol/m^2^) is already filtered to remove pixels with quality assurance values less than 75%. The dataset is available from June 28, 2018 to the present. However, on November 29, 2020, the data version was upgraded to 1.4.0V and this version shows differences in data collection and analysis criteria, for further explanation please see http://www.tropomi.eu/data-products/level-2-products. For this reason the data after November 29, 2020 were excluded from the analysis. For the analysis of the time series, we used the spatial average in a specific region of interest. On the other hand, monthly time averages have been used to show the spatial variability throughout a region of interest. Also, usually there is one observation of NO_2_ per day, and if there are more, an average is taken.

Also, Ozone Monitoring Instrument (OMI/AURA) satellite’s daily tropospheric column concentration measures [53] were retrieved from NASA’s Multi-Decadal Nitrogen Dioxide and Derived Products from Satellites (MINDS) project. OMI images range temporarily from October 2004 to the present. Though, its pixel size is 0.25° × 0.25°, 624 times larger than TROPOMI’s.

Meteorological information was provided by the American National Oceanic and Atmospheric Administration (NOAA) [54]. We obtained the daily average temperature and the daily accumulated precipitation from the studied cities since 2004. In addition, the average daily wind speed in Buenos Aires-Argentina was provided by the Argentinian National Weather Service (SMN for its acronym in Spanish) [55].

The vehicle traffic information is constituted by the number of vehicles per hour that cross the toll booths of Buenos Aires [2] from 2018 to 2020. Raw data present information about the type of vehicle, the toll booth ID and the driving direction among others. This information was aggregated to total vehicle count per day in Buenos Aires. No other city’s circulating vehicle count (or similar) information was freely available or sent upon request from its appropriate ministry.

Population mobility data were taken from the Google Mobility Reports [56], widely used during 2020 [57, 58, 59, 60, 61, 62]. The Google Mobility Reports show changes in baseline daily trips and time spent at home for six categories in 131 countries around the world and their subregions. It establishes the median value of the data collected between January 3^rd^, 2020 and February 6^th^, 2020, and presents data retrieved between February 15^th^, 2020 and December 31^st^, 2020. The data were anonymized and originate from users who agreed to enable location history in their Google accounts [56]. The information used in this study was the percentage variation of the number of visits to residential areas (named residential variation). Their monthly averages are presented in Table S3. Only the information from residential areas was used since it presented the strongest (negative) association with vehicular traffic in Buenos Aires (being the only city from which traffic information was available).

The different sources of data are summarized in Figure 1.

**Figure 1:**
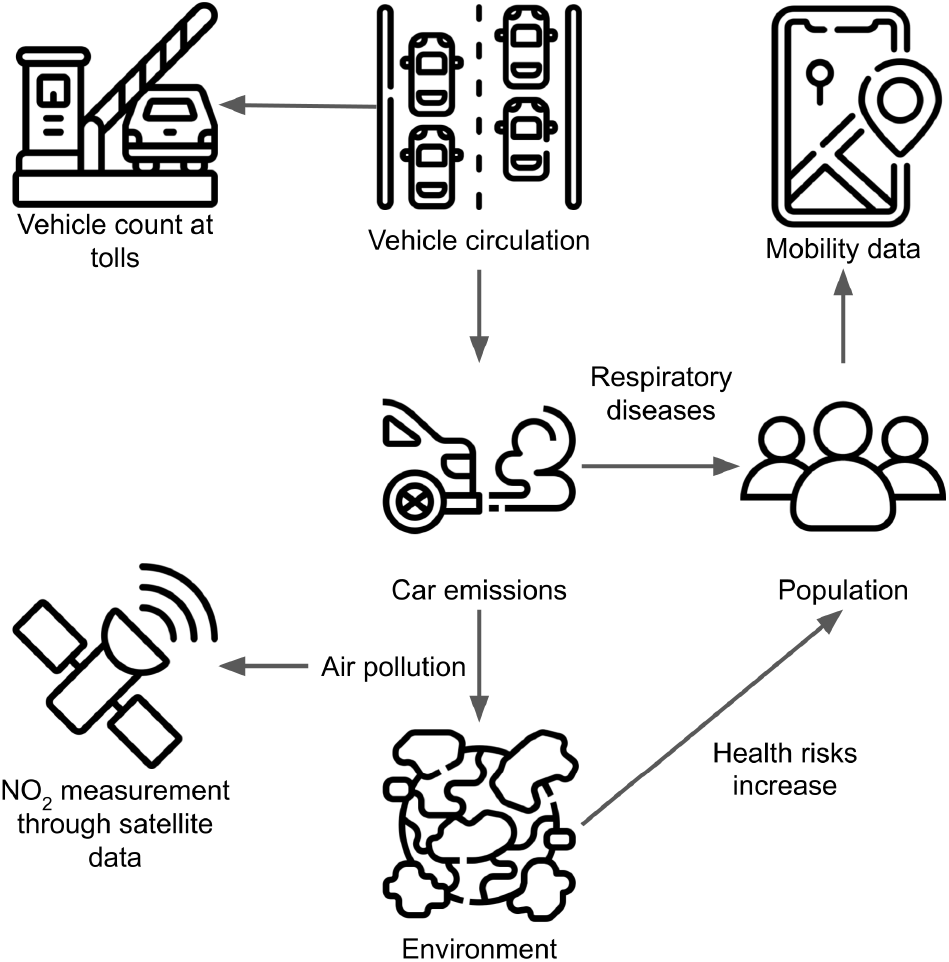
Vehicle circulation is analyzed using vehicle count at tools. Car emissions are monitored from air pollution measures using satellite data. Population mobility is analyzed with cell-phone mobility data.

### 2.2 NO_2_ estimation using satllite data

For each city, a rectangle around boundary delimitation was obtained. From TROPOMI’s images, We selected the pixels whose value was greater than the 70^th^ percentile in all monthly images for each city separately; the extension of these pixels was named the “mask”. To create it, we used the data before the lockdown measures (before march 2020) to avoid possible effects of the quarantine period. Each set of pixels was different for each month. The pixels that were common in all the masks were taken. Each mask was developed in order to measure the reduction of NO_2_ concentration over the urban region of highest emissions. This region differs from the boundary delimitation of the city. On the other hand, given the small amount of OMI pixels covering each city, no mask was applied. The number of TROPOMI’s and OMI’s pixels respectively ranged from 259 and 5 for Buenos Aires to 1986 and 13 for Lima, as it is shown in Figures 2 and S1. From these areas, the average monthly and daily tropospheric NO_2_ concentration average and standard deviation values were calculated (Table S1 and S2). Given the reduced amount of pixels per city for the OMI’s images, only TROPOMI’s tropospheric NO_2_ concentration data was used in this study, except for the historical analysis.

**Figure 2:**
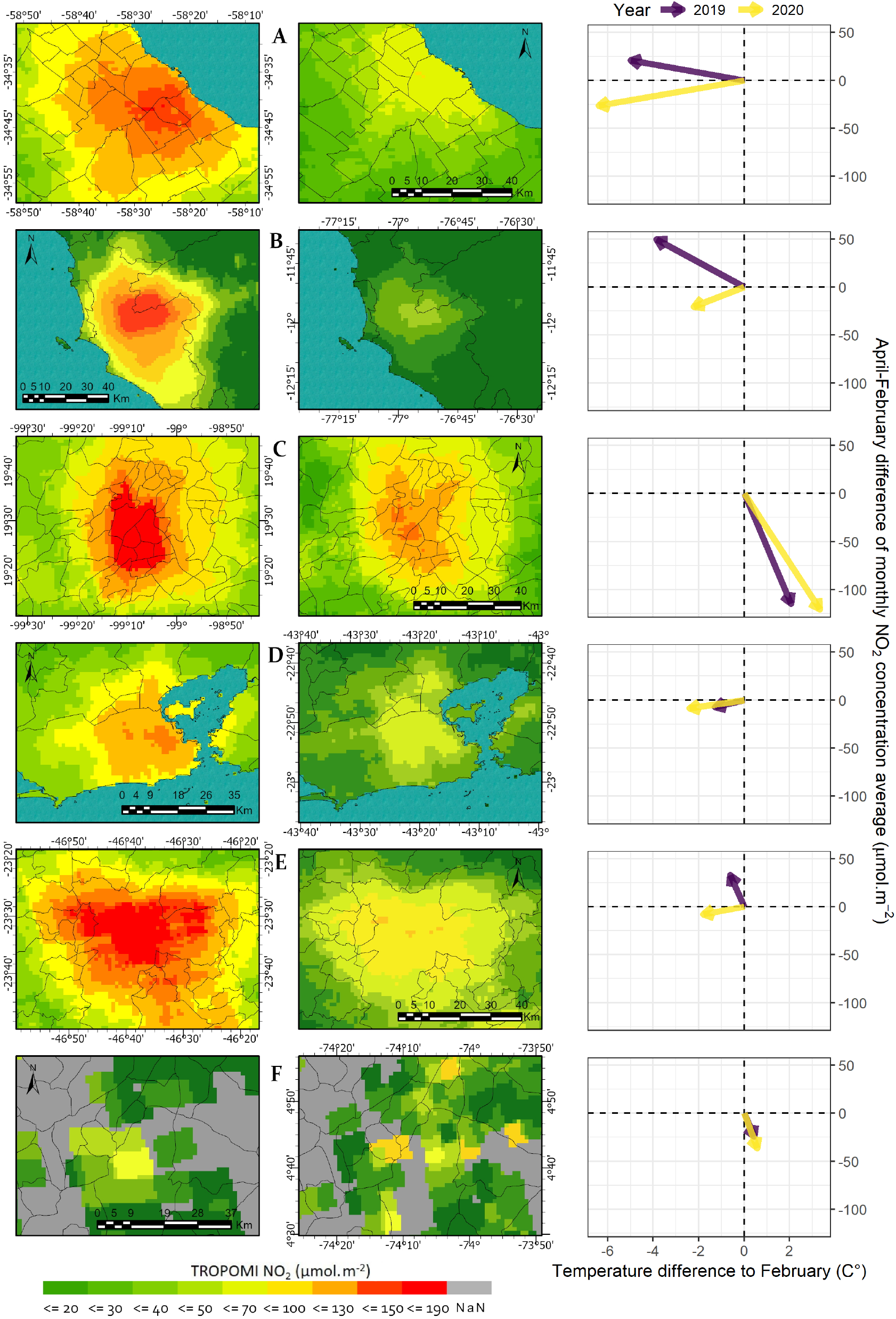
First column: Average tropospheric NO_2_ concentration in April 2019. Second column: Average tropospheric NO_2_ concentration in April 2020. Third column: 2019 and 2020 April-February differences of tropospheric NO_2_ concentration monthly average vs April-February difference of the monthly average temperatures. A) Buenos Aires-Argentina. B) Lima-Perú. C) México DF-México. D) Rio de Janeiro-Brasil. E) São Paulo-Brasil. F) Bogotá-Colombia

### 2.3 Weather and mobility relationship with NO_2_ concentration

Monthly average NO_2_ concentration and temperature for the months of February (before quarantine) and April 2020 (during quarantine) for each city, were taken in order to compare with the monthly average residential variation (Figure 3). We calculated Pearson’s correlation coefficient of the difference between April and February 2020 residential variation and the percentual tropospheric NO_2_ April-February difference scaled by population. We calculated the difference between April 2020 and February 2020 of the tropospheric NO_2_ concentration and the temperature monthly averages (referred to as April-February monthly averages differences) and compared to the ones of 2019, when no traffic restrictions were imposed (Figure 2). Given that TROPOMI satellite was launched in June 2018, previous data doesn’t exist. Because of that and aiming for a better understanding of historical April-February monthly averages differences, we used OMI’s data to reproduce the previous analysis and compared it to the 2004-2019 average (Figure S1)

**Figure 3:**
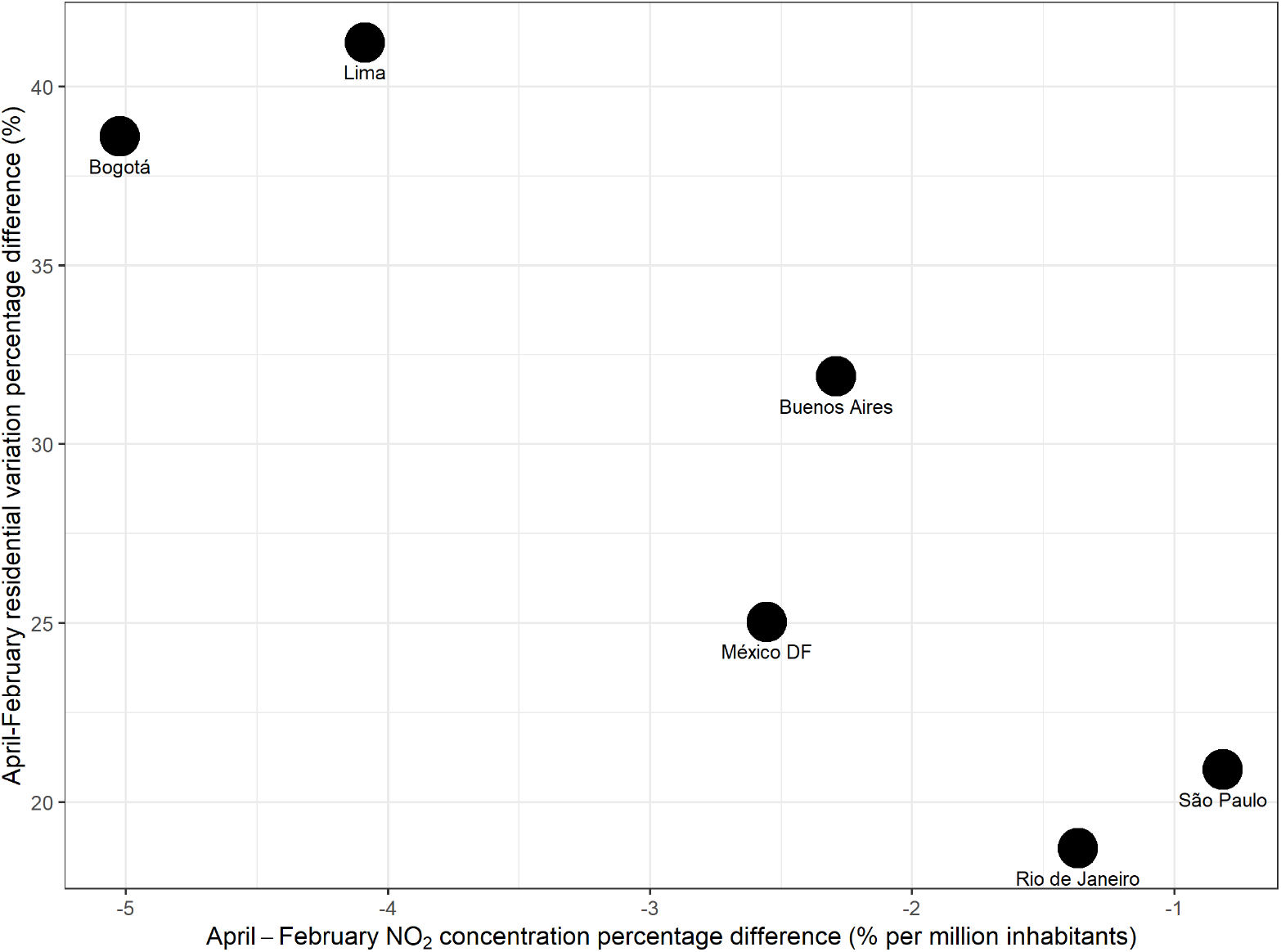
2020 April-February differences for the residential variation and NO_2_ concentration for the six studied metropolitan regions

### 2.4 NO_2_ and mobility model

First, in order to study the strength of the relationship between residential variation and daily vehicle counts, the linear model (1) was constructed:

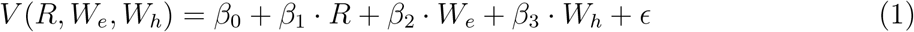

Where *V* is the daily accumulated vehicle count, *R* is the residential variation, *W*_*e*_ are the weekend days, *W*_*h*_ are the holidays, *β*_1_ to *β*_3_ are their respective linear parameters, *β*_0_ is the intercept and *E* is the independent error.

Second, we performed a sensitivity analysis. For this, we built and compared two linear models (2) and (3) which differ only on the variables *V, R, W*_*e*_ and *W*_*h*_ and one additional control model 4 having only the meteorological variables *T*_*lag*2_, *P* and *U*. The comparison was made by calculating the reduction of the residual sum of squares and its significance between models.

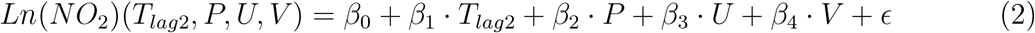

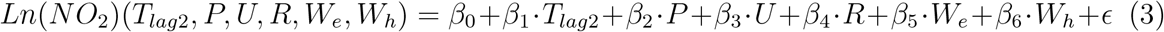

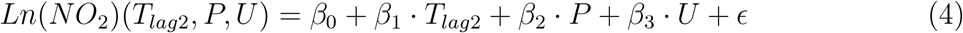

Where *Ln*(*NO*_2_) is the daily tropospheric NO_2_ concentration average in logarithmic scale, *T*_*lag*2_ is the daily temperature average with a two days lag (which improved the models’ fit), *P* is the daily accumulated precipitations, *U* is the daily wind intensity average, *V* is the daily accumulated vehicle count, *β*_1_ to *β*_6_ are the linear parameters, *β*_0_ is the intercept and *ϵ* is the independent error. All assumptions of the linear regression models were satisfied.

## 3 Results

We have observed a decrease in tropospheric NO_2_ concentration in every city during almost every month between March to December 2020 compared to the year before. The stronger decreases were seen in April. The decrease was 21.3% in Mexico DF, 28.94% in Bogotá, 39.1% in Rio de Janeiro, 39.3% in São Paulo, 56.3% in Buenos Aires, and 71.8% in Lima (Figure 2, first and second column). These results are in agreement with previous researches: Represa et al. (2020) [20] observed a 54% reduction of the tropospheric NO_2_ mean column concentration during 2020 in Buenos Aires. In México DF, a 18.1% reduction was observed in 2020 compared to the 2017-2019 period [63]. In Bogotá, a 20% tropospheric NO_2_ concentration reduction was observed during the lockdown [64]. In Rio de Janeiro and São Paulo, a 9.1%-41.8% reduction was measured in different locations of the cities [65]. Lima was pointed out as one of the strongest decreases worldwide [66, 67] with a 61% NO_2_ concentration reduction in April 2020 compared to 2019 [68, 67].

The April-February tropospheric NO_2_ concentration and temperature monthly averages differences showed a change between 2019 and 2020 (Figure 2, third column). As expected by seasonality, the April-February temperature monthly average difference is negative for the southern hemisphere cities (Buenos Aires, Lima, Rio de Janeiro and São Paulo), and positive for the northern hemisphere cities (México DF and Bogotá). Even though tropospheric NO_2_ concentration is inversely related to the air temperature (as observed in almost every city of this study between 2004 and 2019 as seen in Figure S1, third column), this relationship was not widely observed in 2020: Buenos Aires, Lima and São Paulo showed less tropospheric NO_2_ concentration in April than in February, yet the temperature was higher in February. In addition, Bogotá showed a higher NO_2_ decrease in 2020 compared to 2019 despite a very similar April-February temperature difference. Nevertheless, Mexico DF showed a slight higher decrease in 2020 compared to 2019 on the April-February tropospheric NO_2_ concentration difference as expected by the higher temperature difference in 2020. On the other hand, Rio de Janeiro showed both in 2019 and 2020, less tropospheric NO_2_ concentration in February than in April, being February always warmer. OMI’s historical data support these affirmations. Nevertheless, for the Brazilian cities, the 2020 April-February difference was similar to the historical 2004-2019 observations average. Several factors may play an important role in this observation: On one hand, Brazil presented a less restrictive lockdown compared to the other studied countries [18, 19]. On the other hand it was not possible to apply a mask over the reduced amount of pixels in OMI’s images (as seen for the other cities, compared to TROPOMI’s data). Conjointly, these factors may hamper differences detection between 2020 and historical data.

The 2020 April-February residential variation difference and the tropospheric NO_2_ concentration percentage difference (Figure 3) showed an interesting ordering of the cities: the ones with the less restrictives lockdowns (Rio de Janeiro, São Paulo and Mexico) have the smallest residential variation and absolute NO_2_ concentration percentage differences, contrary to the cities with more restrictive lockdowns (Lima and Bogotá). Similarly to Li (2020) [59] in Singapore and Wong (2021) [46] in the USA, we have also observed a linear correlation between tropospheric NO_2_ concentration and another population mobility indicator. Our results showed that the strongest NO_2_ reduction was seen in Lima (Perú), likewise Corpus-Mendoza (2021) [48] who observed the strongest decrease in NO_2_ AQI in Latin America being in Lima, and the second strongest globally.

Importantly, when we compared the different cities there was a strong relationship between the April-February residential variation and the population-scaled tropospheric NO_2_ concentration percentage differences. Hence, we observed a significant negative linear correlation (*ρ* = -0.90, p-value = 0.01) between those variables (3). As for the relationship between the daily residential variation and daily accumulated vehicle count in Buenos Aires, model (1) showed a strong linear association 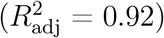 indicating a great collinearity and information shared between these features. In addition, models (2) to (4) were satisfactorily adjusted. Model (4) was proven significantly different (p-value < 0.001) to models (2) and (3), both not significantly different from each other. These results are reflected on the similarity of the models’ estimated parameters’ confidence intervals shown in Table 1 as for the temperature and wind, their related estimated confidence intervals consistently overlap in models (2) and (3) contrary to the temperatures’ estimated parameter confidence interval in model (4). This sensibility analysis allowed us to conclude that toll’s vehicle count data and residential variation data in Buenos Aires shared a large amount of information in order to explain the logarithmic tropospheric NO_2_ concentration behavior during the 2020 COVID-19 lockdown.

**Table 1:** Model estimates’ confidence intervals. *V* : daily accumulated vehicle count, *T*_*lag*2_: daily temperature average with a two days lag, *W*_*h*_: holidays, *W*_*e*_: weekend days, *U* : daily wind speed average, *R*: residential variation

| | Intercept | $V$ | $T_{lag2}$ | $W_h$ | $W_e$ | $U$ | $R$ |
| --- | --- | --- | --- | --- | --- | --- | --- |
| Model (2) | [-9;-8.6] | [2.6e-06;3.7e-06] | [-0.056;-0.034] |  |  | [-0.11;-0.075] |  |
| Model (3) | [-7.8;-7.2] |  | [-0.055;-0.032] | [-0.7;-0.28] | [-0.63;-0.39] | [-0.11;-0.073] | [-0.028;-0.015] |
| Model (4) | [-8.8;-8.3] |  | [-0.031;-0.0079] |  |  | [-0.11;-0.071] |  |

These findings are meaningful because understanding NO_2_ dynamics and its relation with human activities is one of the first steps to reduce its local and global concentration. It may help to better comprehend its direct and indirect impacts on the urban population’s health and life quality and on the environment. NO_2_ is a mainly anthropogenic gas that affects population health. Our results show how drastically and rapidly tropospheric NO_2_ concentrations are related to the variation of urban population mobility in the largest cities of Latin America which was not described before. Venter et. al. 2020 [69] had previously studied the Google’s mobility report work indicator and an Apple’s vehicle driving indicator relationship with tropospheric NO_2_ concentration during COVID lockdowns in, mostly, northern developed countries. Differently, our study aims to compare the equivalence of Google’s mobility data reports and in-city direct vehicular count over the prediction of tropospheric NO_2_ concentration. Our findings show no significant differences using one or the other variable at the tropospheric NO_2_ concentration prediction. Additionally, the tropospheric NO_2_ concentration and population mobility decreased during the lockdown period in Latin America indicating a correlation between the decrease in NO_2_ concentrations and human mobility, in agreement to Venter et. al.

Nevertheless, there are some limitations around these conclusions. Firstly, satellite data used were downloaded as L3 which are interpolated values from a L2 rawer data set. Secondly, during the study period, Bogotá was affected by the persistent presence of clouds. When GEE transforms L2 data into L3, filtering with qa_value is applied. Pixels with missing values correspond to areas where the tropospheric column retrieval has a larger uncertainty. Consequently, the size of the data used to draw conclusions over Bogotá was small and the results are not as reliable as for the other cities. Those meteorological conditions were confirmed on the worldview platform [70]. Thirdly, due to the change in the version of the TROPOMI data, it was not possible to carry out a study on the recovery of NO_2_ concentration levels with the flexibility of mobility restriction measures. As there was usually one observation of NO_2_ per day, these values were taken as the representatives of their respective days, this represented a source of error since it didn’t take into account the daily variability present in NO_2_ concentrations. Lastly, Google Mobility Reports’ spatial precision did not exactly correlate with the satellite data spatial extension due to an unclear territorial delimitation on the mobility data and the fact that the area of highest NO_2_ concentration was not restricted by boundary delimitation.

## 4 Conclusion

The results of this study confirm our initial hypothesis; they showed that the tropospheric NO_2_ concentration and population mobility decrease during the lockdown period in Latin America are correlated. This study is a background for public policy decision makers. The relationship between mobility and air quality could be used as an input for urban planification and for climate change mitigation measures. In addition to the results, our work provides methodological strategies for data processing.

Also, the cities that showed the strongest decrease of NO_2_ concentration and to the population mobility reduction during April 2020 (Lima and Buenos Aires), seemed to be the cities with the most restrictive lockdowns. In contrast, the cities with the weakest decrease of NO_2_ concentration and population mobility (México, Rio de Janeiro and São Paulo) would appear to be the ones with the least restrictive measures. Nevertheless, this study did not address the degree of intensity of these restrictive measures but, it’s a precedent for future works focusing on this topic.

Although this work, and all the previously mentioned, had shown an important relation between NO_2_ concentration and population mobility, causation is harder to prove being observational studies. Though it is possible that the globally observed NO_2_ concentration decrease during the lockdowns is not caused by the population mobility reduction, it is very unlikely not to be, and the more studies like ours there are, the more this hypothesis is reliable. We proposed simple methods, as linear regressions, and they showed a great fit. These results highlight the unquestionable correlation between these features.

## 5 Data accessibility

Data and scripts are available on github (https://github.com/matiaspoullain/NO2-pollutiondecrease-Latin-America-COVID-repository)

## Data Availability

All data produced are available online at
https://github.com/matiaspoullain/NO2-pollution-decrease-Latin-America-COVID-repository

## 6 Acknowledgement

Authors thank Fernanda Garcia for fruitful discussion.

## 10 Supplementary materials

**Table S1:** Observed averages and mean standard error of the TROPOMI’s monthly torpospheric NO_2_ concentration (*µ*mol.m^-2^)

| Month | Bogotá<br>(n=367) | Buenos Aires<br>(n=259) | Lima<br>(n=1986) | México DF<br>(n=1165) | Rio de Janeiro<br>(n=750) | São Paulo<br>(n=633) |
| --- | --- | --- | --- | --- | --- | --- |
| Feb | 61.87 (9.09) | 84.62 (7.10) | 46.31 (4.92) | 216.26 (19.41) | 52.96 (5.37) | 77.62 (4.69) |
| Mar | 38.36 (6.30) | 87.20 (7.91) | 38.57 (4.05) | 185.19 (15.90) | 40.04 (3.67) | 70.87 (4.18) |
| Apr | 25.50 (1.78) | 58.17 (3.88) | 25.86 (2.35) | 95.67 (5.06) | 44.34 (3.60) | 69.82 (5.15) |
| May | 26.91 (1.16) | 88.32 (6.76) | 47.95 (5.16) | 86.28 (5.50) | 47.29 (4.05) | 101.96 (7.36) |
| Jun | 20.99 (1.97) | 136.70 (9.03) | 78.82 (8.12) | 95.79 (5.97) | 77.47 (6.08) | 110.85 (8.36) |
| Jul | 16.35 (1.23) | 133.90 (8.82) | 103.97 (9.89) | 109.96 (9.87) | 75.15 (7.89) | 151.32 (10.32) |
| Aug | 31.95 (1.91) | 83.19 (5.85) | 99.73 (9.30) | 87.92 (10.70) | 64.96 (5.45) | 150.73 (9.48) |
| Sep | 28.74 (2.51) | 117.70 (12.30) | 92.94 (7.96) | 99.95 (11.89) | 66.03 (5.59) | 117.41 (6.91) |
| Oct | 21.42 (3.46) | 105.02 (8.17) | 83.07 (7.07) | 167.31 (13.92) | 53.70 (4.49) | 99.50 (5.29) |

**Table S2:** Observed averages and mean standard error of the OMI’s monthly torpospheric NO_2_ concentration (*µ*mol.m^-2^)

| Month | Bogotá | Buenos Aires | Lima | México DF | Rio de Janeiro | São Paulo |
| --- | --- | --- | --- | --- | --- | --- |
| Feb | 30.30 (3.26) | 57.54 (5.19) | 22.05 (1.64) | 103.65 (8.83) | 29.71 (2.68) | 32.74 (3.45) |
| Mar | 29.39 (4.04) | 38.39 (3.81) | 17.84 (1.81) | 85.46 (7.72) | 42.81 (3.64) | 38.86 (2.83) |
| Apr | 21.53 (0.92) | 53.31 (10.12) | 15.61 (1.16) | 44.45 (3.58) | 33.95 (3.10) | 48.45 (3.94) |
| May | 11.13 (0.45) | 73.28 (8.54) | 19.29 (1.73) | 55.04 (3.93) | 41.27 (2.95) | 55.68 (3.73) |
| Jun | 19.70 (2.00) | 110.26 (13.59) | 26.55 (1.79) | 47.73 (4.08) | 47.56 (2.62) | 64.22 (3.70) |
| Jul | 17.44 (1.52) | 89.60 (10.30) | 31.58 (2.46) | 53.13 (2.79) | 48.34 (3.66) | 73.39 (6.72) |
| Aug | 17.03 (2.60) | 63.86 (6.48) | 29.37 (2.31) | 45.37 (3.11) | 42.20 (3.90) | 81.06 (6.93) |
| Sep | 18.84 (2.54) | 83.85 (8.51) | 25.84 (1.75) | 53.16 (4.30) | 50.70 (3.17) | 64.22 (4.25) |
| Oct | 17.37 (1.96) | 69.57 (5.58) | 26.35 (1.59) | 69.85 (5.57) | 45.29 (3.48) | 52.36 (4.07) |
| Nov | 18.23 (2.19) | 43.76 (3.83) | 22.62 (1.45) | 89.76 (7.83) | 27.26 (2.83) | 38.94 (3.26) |
| Dec | 11.48 (2.64) | 52.03 (3.93) | 21.86 (2.69) | 101.59 (8.74) | 36.92 (2.57) | 43.98 (3.62) |

**Figure S1:**
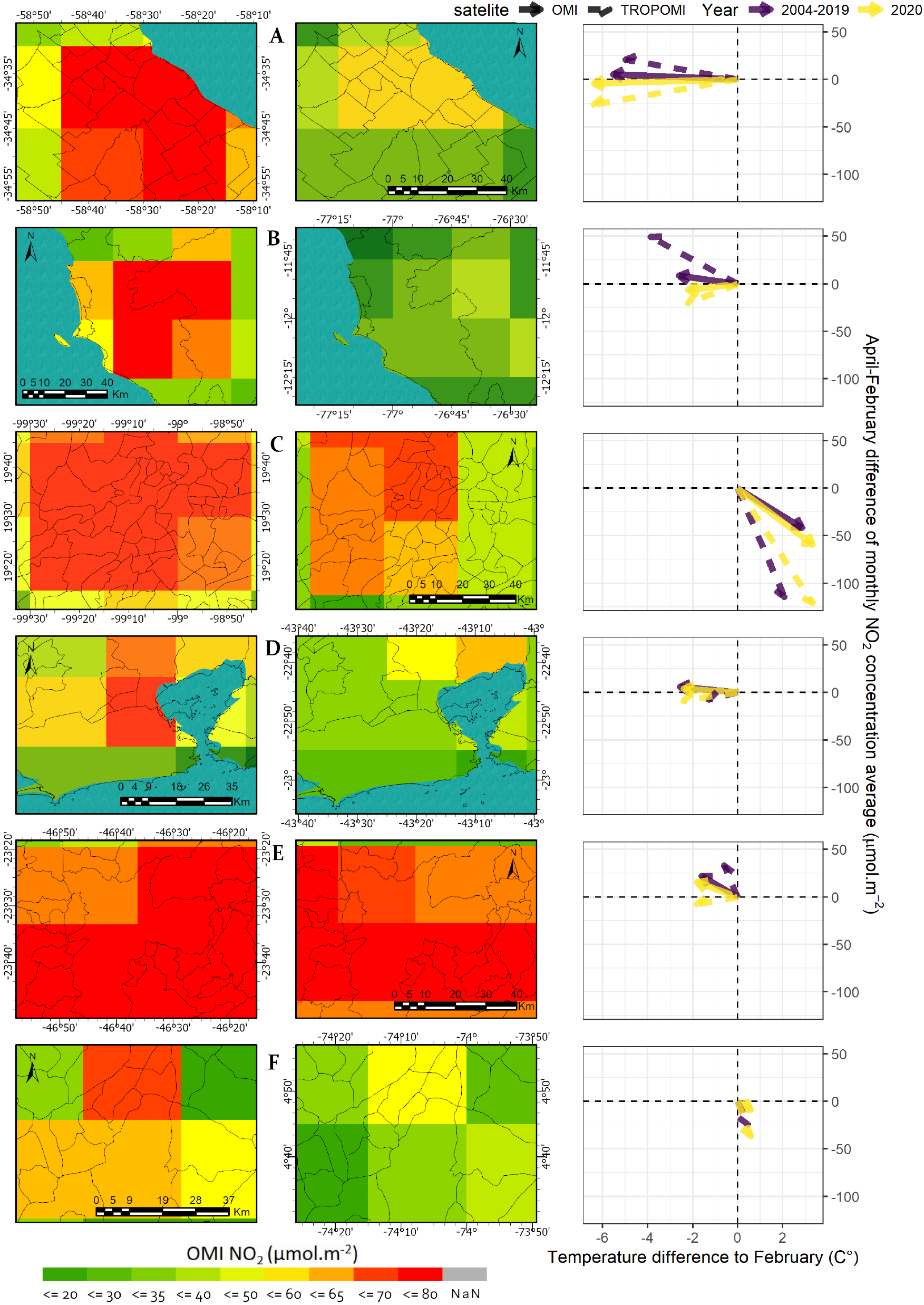
First column: Average OMI’s tropospheric NO_2_ concentration in April 2019. Second column: Average OMI’s tropospheric NO_2_ concentration in April 2020. Third column: 2004-2019 average and 2020 April-February differences of tropospheric NO_2_ concentration monthly average vs April-February difference of the monthly average temperatures for OMI and TROPOMI’s measurements. A) Buenos Aires-Argentina. B) Lima-Perú. C) México DF-México. D) Rio de Janeiro-Brasil. E) São Paulo-Brasil. F) Bogotá-Colombia

**Table S3:** Observed averages and mean standard error of the monthly residential variation (%)

| Month | Bogotá | Buenos Aires | Lima | México DF | Rio de Janeiro | São Paulo |
| --- | --- | --- | --- | --- | --- | --- |
| Feb | -1.13 (0.10) | 1.47 (0.25) | -0.07 (0.03) | -1.13 (0.03) | 2.20 (0.30) | 1.60 (0.35) |
| Mar | 15.87 (0.63) | 16.65 (0.54) | 21.19 (0.65) | 7.10 (0.27) | 9.81 (0.33) | 9.42 (0.37) |
| Apr | 37.47 (0.20) | 33.37 (0.11) | 41.17 (0.16) | 23.90 (0.17) | 20.90 (0.10) | 22.50 (0.14) |
| May | 30.39 (0.19) | 29.35 (0.11) | 36.84 (0.13) | 25.19 (0.18) | 19.61 (0.10) | 21.00 (0.14) |
| Jun | 26.10 (0.18) | 27.37 (0.07) | 31.17 (0.11) | 22.33 (0.16) | 16.40 (0.09) | 18.30 (0.10) |
| Jul | 25.74 (0.16) | 28.00 (0.10) | 24.94 (0.11) | 18.81 (0.14) | 12.94 (0.09) | 15.26 (0.10) |
| Aug | 24.55 (0.18) | 24.13 (0.08) | 23.48 (0.13) | 15.68 (0.16) | 10.55 (0.09) | 13.10 (0.11) |
| Sep | 18.50 (0.17) | 21.70 (0.09) | 22.03 (0.12) | 15.53 (0.15) | 9.83 (0.10) | 11.63 (0.12) |
| Oct | 15.35 (0.17) | 19.19 (0.11) | 18.90 (0.09) | 14.23 (0.15) | 9.13 (0.06) | 10.32 (0.10) |
| Nov | 14.27 (0.18) | 15.70 (0.10) | 17.43 (0.09) | 14.03 (0.17) | 8.87 (0.12) | 9.60 (0.13) |

